# Heat Stress in relation to Sleep Health among Farmers

**DOI:** 10.1101/2025.07.28.25332331

**Authors:** Wensu Zhou, Symielle A. Gaston, Bethany T. Ogbenna, Christine G. Parks, Dale P. Sandler, Chandra L. Jackson

## Abstract

**Background:** While high temperatures are linked to poor sleep, few studies considered heat stress, especially among outdoor workers.

**Objective:** We investigated associations between heat stress and sleep health among farmers.

**Methods:** The study included 8,203 male participants from Iowa (78%) and North Carolina (NC, 22%), enrolled in the Agricultural Health Study (2013–2015), who were actively farming when data were collected. Survey data on sleep, demographics and covariates were linked to daily wet bulb globe temperatures (WBGT) from May 2013–September 2015. We calculated absolute heat stress by averaging WBGT over 2, 5, and 7 days before the interview. Relative heat stress (i.e., the difference between absolute heat stress and the 92.5th percentile of WBGT) was also calculated. WBGT was categorized by heat stress risk (low, moderate, high). Sleep measures included short nightly sleep duration (<7 hours), daytime sleepiness (≥3 days/week), daytime napping (yes), and long napping duration (≥30 minutes). Poisson regression with robust variance was used to estimate sociodemographic-adjusted prevalence ratios and 95% confidence intervals (PR[CI]) in Iowa and NC, separately.

**Results:** Farmers had a mean age of 63 years [SD=10.1]; 37.8% reported short sleep, 8.1% daytime sleepiness, 44.6% daytime napping, and 17.1% long naps. Mean absolute WBGT were 70.4°F (SD=6.36) in Iowa and 77.7°F (SD=7.83) in NC. In Iowa, moderate heat stress (2-day average) was associated with higher short sleep prevalence (PR= 1.04 [1.00–1.07]). In NC, higher absolute and relative WBGT (2-/5-/7-day average), as well as moderate (2-/7-day) and high (2-day) heat stress were associated with daytime napping (e.g., PR _2-day absolute WBGT_= 1.02 [1.01-1.04]). In both states, high heat stress was associated with a lower prevalence of long naps (e.g., PR_Iowa, 2-day heat stress_= 0.86 [0.83-0.89]).

**Significance:** Among farmers, heat stress was associated self-reported indicators of poor sleep. Future research with objective sleep measures is warranted.

**Impact Statement:** This study is among the first to determine associations between daytime heat stress exposure, measured by wet bulb globe temperature (WBGT), with both nighttime sleep and daytime napping among farmers who reside on farms and are actively engaged in farming. Our results suggest that heat stress may be related to short nighttime sleep duration and daytime napping but shorter nap duration. These findings have important implications for improving sleep health, which contributes to favorable outcomes such as injury prevention in agricultural workers.

## INTRODUCTION

Sleep disturbances are prevalent in the general US population. For example, a recent nationally representative study estimated that the proportion of adults in the United States (US) with short sleep (<7 hours) was 23.1%, 29.8% reported trouble sleeping, and 27.2% reported daytime sleepiness ^1^. Moreover, trouble sleeping was more prevalent among older adults compared to younger individuals ^1^, which is important because the number of individuals aged 65 years or older is expected to increase by 47% by 2050 ^2^. Sleep disturbances are of great public health concern due to their high prevalence and associations with many chronic diseases and mental health conditions ^3^. Therefore, there is a critical need to identify and address factors that impede optimal sleep ^4^.

Numerous studies have linked high daytime temperatures with increased risk of heat-related illnesses ^5–10^ as well as with disruption of essential physiological processes, including those related to sleep duration and quality ^11–16^. Specifically, epidemiological studies using mean or maximum daytime ambient temperature have found associations between heat exposure and both reduced sleep duration and poorer sleep quality among samples of the general population ^15, 17, 18^. Beyond absolute temperature, relative high temperature—which considers both the intensity and duration of high temperatures—is also important. Currently, only one study of older adults has investigated relative heat (defined by temperature threshold combined with duration [days]) in relation to sleep, and found that exposure (e.g., daily minimum temperature above the 92.5th percentile for at least three days) was associated with shorter sleep duration ^19^. Therefore, little is known about the relationship between relative daytime temperature and different aspects of sleep, including sleep disturbances.

However, relying solely on ambient temperature may not accurately reflect the heat burden experienced by the human body. For example, high humidity can impair the body’s ability to dissipate heat through sweating, thereby increasing heat burden ^20^. When the body cannot effectively release excess heat, it experiences heat stress—a condition marked by physical discomfort and physiological strain due to exposure to a hot environment, typically characterized by an increase in core body temperature and heart rate ^21^. By combining temperature and relative humidity, the Heat Index (HI) serves as a biometeorological indicator that reflects heat stress on the human body and has been used in previous studies ^22, 23^. However, other environmental factors—such as wind speed—also influence the body’s heat balance and can further modify heat stress levels ^24^. Thus, assessing heat exposure in relation to sleep health should involve indicators that more accurately reflect heat stress ^25^. The Wet Bulb Globe Temperature (WBGT) ^26^—which incorporates wind speed, cloud cover, humidity, and sun angle—offers a more comprehensive measure of heat stress and is better suited for evaluating thermal conditions during hot-weather activities than ambient temperature alone ^27, 28^. Several studies investigating the relationship between heat stress and sleep in the general population used WBGT and found associations between heat stress and sleep disturbances; yet, these studies often measured sleep duration alone rather than multiple dimensions of sleep health, involved relatively small sample sizes, and yielded mixed findings ^28–32^. Moreover, the impact of heat exposure on health outcomes varies depending on occupation and working conditions ^33^; however, prior research focused primarily on populations who are not outdoor workers and are, therefore, less likely to be occupationally exposed to daytime heat. Further research of sleep health among populations with relatively higher heat stress exposure burdens is warranted.

Outdoor workers, particularly those engaging in strenuous physical activity like agricultural workers, are more affected by heat-related health outcomes relative to the general population ^34^. During farming production periods, which often overlap with the hottest months, farmers are more likely to experience elevated body core temperature and dehydration due to prolonged exposure to solar radiation ^35^. Elevated core body temperature can contribute to discomfort, and heat exposure may impair thermoregulation by skin vasodilation as well as contribute to increased wakefulness, thereby impacting sleep ^36^. Thus, it is plausible that heat exposure is associated with loss of sleep duration and sleep disturbances. Moreover, intensive outdoor labor also makes workers more likely to have fatigue and sleep disturbances ^34, 37^, which has implications for other outcomes, such as work injures ^38^. There is limited empirical evidence examining the association between heat stress exposure and sleep among farmers. Additionally, published literature indicates that age modified the relationship between heat exposure and sleep: as daily mean temperatures rose, sleep duration declined — especially among older adults (aged over 65 years) compared to younger age groups (19 to 65 years) ^39^. Since older people are more vulnerable to heat primarily due to age-related diminished thermoregulatory function and the presence of comorbidities ^40^, it is also important to consider age as a potential effect modifier. But, to our knowledge, studies to date have yet to compare the associations between heat stress and sleep between older and younger farmers.

To address these research gaps, the objective of this study was to investigate both absolute and relative WBGT in relation to sleep duration and disturbances among farmers in the US. In addition, we investigated age as a potential effect modifier of the relationship between heat stress and sleep. We hypothesized that heat stress is associated with short sleep duration, daytime sleepiness, daytime napping, and long napping duration. Further, we hypothesized that these associations are stronger among older than younger farmers.

## METHODS

### Data source and study population

We used data (Releases: P1REL202210.00) from the Agricultural Health Study (AHS), a prospective cohort study of 89,655 licensed pesticide applicators (mostly farmers) and their spouses from Iowa and North Carolina (NC), enrolled in 1993-1997. Participants completed self-administered questionnaires. Data about lifetime pesticide use, and demographic and medical history were collected. Participants provided implied consent at enrollment and active consent during follow-up surveys following protocols approved by the relevant Institutional Review Boards. Detailed information on the AHS has been previously published ^41^. Since enrollment, four follow-up surveys were conducted in 1999-2003, 2005-2010, 2013-2015, and 2019-2021.

This study mainly used cross-sectional data from the 2013-2015 survey, which collected comprehensive data on sleep.

A total of 42,331 participants completed the survey. The study sample excluded farmers’ spouses (n=18,186) and female farmers (n= 661), due to potential differences in heat exposure between male farmers and their spouses and the small sample size of female farmers, which limited power to conduct sex-specific analyses. Among the remaining 23,484 male participants, the study sample excluded those who did not reside on a farm or personally perform farm work in the past 12 months (n=10,669), were not interviewed during the warm season months of May to September (n=4,040) ^42^, were missing key covariate information and sleep data (n=526), or were WBGT data (n=46). Therefore, a total of 15,281 of participants were ineligible, resulting in a total analytic sample of 8,203 farmers (n= 6,403 [Iowa]; n=1,800 [NC]) with complete data. A detailed flowchart of the sample selection is presented in Figure S1. Excluded participants were generally older than those included, with a higher proportion over age 70, but the distribution of education and marital status was similar (Table S1).

### Exposure assessment: Heat stress

Iowa is located in the central region of the US and has a four-season climate. The average temperature during the warm season months (May to September) from 2013 to 2015 ranged from 66.4°F (19.1°C) in 2014 to 67.8°F (19.9°C) in 2013, and 67.7°F (19.8°C) in 2015 ^43^. The weather in NC, located along the Atlantic coast, is humid with very warm summers and moderately cold winters. During the warm season months (May to September) from 2013 to 2015, average temperatures in North Carolina ranged from 72.0°F (22.2°C) in 2013 to 73.0°F (22.8°C) in 2014, and 74.4°F (23.6°C) in 2015 ^43^.

The existing heat data linked to the AHS are derived from satellite-based climate data provided by the North American Regional Reanalysis (NARR), a product of the National Centers for Environmental Prediction (NCEP) and the National Oceanic and Atmospheric Administration (NOAA). These data are publicly accessible and available on a daily basis (https://www.ncei.noaa.gov/products/weather-climate-models/north-american-regional). NARR data are provided at a 32-kilometer resolution, and geocoded baseline participant home addresses were matched to the nearest grid cell. Thus, temperature and Heat Index (HI) values were unique to each grid cell that is linked to a participant’s residential address. These HI values were then used to calculate the Wet Bulb Globe Temperature (WBGT) from 2013 to 2015, using the formula WBGT = −0.0034 HI^2^ + 0.96 HI - 34 ^44^. The participants’ residential addresses at enrollment were geocoded and linked to the WBGT data. Since sleep is acutely affected by temperature ^11^, short-term heat exposure is the primary focus of the present study.

The critical exposure window periods were defined by the following number of days prior to the interview date: 2-day, 5-day, and 7-day moving averages. For example, a 2-day moving average was defined as the two-day average levels of WBGT exposure prior to interview day.

We calculated relative exposure to WBGT to represent the intensity of heat exposure. This approach defines exposure using several percentile-based thresholds of the WBGT distribution over a specified period, and these thresholds are based on long-term, location-specific temperature records, allowing for a more accurate estimation of heat-related health impacts ^45^. In previous studies that aimed to capture relative heat exposure (e.g., using daily temperature), commonly used thresholds included the 90th, 92.5th, 95th, and 97.5th percentiles ^46, 47^ Consistent with prior literature, we calculated the 90th, 92.5th and 95th percentiles for the WBGT series in the warm season (May to September) of 2013-2015 for each participant based on their locations ^19, 46^. We considered the 92.5th percentile as the main threshold to determine relative heat stress; this level reflected mild to moderate heat intensity that could directly increase health risks ^48^. Relative exposure was then defined as the difference between absolute WBGT and the corresponding percentile threshold.

We created categorical exposure measurements based on a real-world heat safety guide developed for outdoor athletic purposes ^42^. This guide helps prevent heat-related illnesses and injuries by defining WBGT thresholds that capture risk of heat stress for each heat safety region, considering local long-term climate conditions, thus providing practical recommendations for physical activity. The thresholds were previously determined by U.S. region (n=3) based on geographic distribution, with variations reflecting local climate adaptation ^42^. This use of these thresholds has been endorsed in the Expert Consensus Statement on Exertional Heat Illness: Recognition, Management, and Return to Activity ^49^. Based on the guidelines, the three heat stress exposure categories were: ‘normal activity’ (noted as ‘low risk’ in the present study; WBGT <78.8 °F for Iowa and <82.1 °F for NC), ‘plan intense or prolonged activity with discretion’ (moderate risk; WBGT 78.8-83.7 °F for Iowa and 82.1-86.0 °F for NC), and ‘limit or cancel outdoor activity’ (high risk; WBGT >83.7 °F for Iowa and >86.0 °F for NC).

### Outcome assessment: Sleep duration, daytime sleepiness, and napping

In the present study, the following sleep measures reflecting dimensions of sleep health were included ^50^. Nightly sleep duration was determined based on responses to “How many hours of sleep do you get each night?”. We dichotomized responses as <7 hours vs. ≥7 hours, consistent with the joint recommendations of the American Academy of Sleep Medicine and the Sleep Research Society ^51^. We defined daytime sleepiness based on responses to the question “How often do you feel sleepy most of the day?”, categorized as ≥3 days/week vs. <3 days/week. Daytime sleepiness was used as a proxy for poor sleep quality, as its symptoms— such as untimely and unwanted sleep episodes—are indicative of poor sleep quality ^52^. Two other measures, daytime napping (“Do you nap during the day?”, yes vs. no) and duration of napping among participants who reported napping (“How long do you nap?”, ≥30 minutes vs. <30 minutes) were also included.

### Potential effect modifier

Age was considered a potential effect modifier given prior evidence that identifies older adults as a vulnerable population ^53^. Participants’ ages were categorized into two groups: <60 years and ≥60 years, based on the United Nations definition of an older person as someone aged 60 or above ^54^ and supported by a sufficiently balanced age distribution in our dataset.

### Potential confounders

Potential confounders were identified based on a directed acyclic graph (DAG). Our approach aligns with epidemiologic recommendations to reduce overadjustment which may bias the measure of association ^55^. According to the DAG (Figure S2), current age and marital status (both in the 2013-2015 survey), and race and ethnicity, and educational attainment (collected at enrollment) were considered potential confounders in the analysis.

### Statistical analysis

We conducted cross-sectional analyses in Iowa and NC, separately, primarily to account for the substantial geographic differences between the two states. Descriptive statistics of demographic information and sleep characteristics were estimated as means and standard deviations (SDs) for continuous variables and counts and percentages for categorical variables. Using t-tests or chi-square tests, we compared the distributions of all variables in Iowa and NC.

We estimated the relationship between heat stress (i.e., absolute WBGT, relative WBGT, and heat stress categories) and measures of sleep health, by state. Using Poisson regression with robust variance estimation, we calculated prevalence ratios (PR) and 95% confidence intervals (CIs). For absolute WBGT and relative WBGT, we assessed associations with a 1 SD increase. For heat stress categories (i.e., low, moderate, and high risk), the low-risk category (i.e., ‘normal activity’) served as the reference. All models were adjusted for age, race and ethnicity, marital status, and educational attainment. We evaluated effect modification by age in stratified models and by testing cross-product interaction terms.

We conducted sensitivity analyses to determine whether our study design affected the findings. First, we included female farmers with complete demographic and WBGT data in the analysis (n = 96 for Iowa and n = 54 for NC). Next, we modified the short sleep duration cut point to <8 hours versus ≥8 hours since sleep duration is often misreported/overestimated ^56, 57^. There is no consensus about napping duration recommendations ^58^, so we used a different cut point for napping duration (i.e., ≥1 hour versus <1 hour), similar to a prior study ^59^. Third, we modified the thresholds for relative heat stress measurements by using the 90th and 95th percentiles of WBGT. This was employed to assess whether using lower or higher thresholds than those in the main analysis would alter the associations between WBGT and measures of sleep health. We also sought to determine whether the association between heat stress and sleep were further influenced by different levels of relative heat stress. Lastly, we adjusted for whether participants took naps in the models for sleep duration, and for sleep duration in the models for daytime napping (yes vs. no, not napping duration since that is only captured among the people who reporting taking naps).

## RESULTS

### Study population characteristics

Table 1 presents the participant sociodemographic characteristics, heat stress measures, and sleep outcomes. Among the 8,203 farmers, 78.1% (n=6,403) were from Iowa and 21.9% (n=1,800) from NC, with a mean age of 62.5 years (SD=9.84) in Iowa and 64.8 years (SD=10.6) in NC. Almost all participants were White (99.1%), while the remaining participants identified as another race (0.9%). Most completed high school or more education (95.8%), with a higher proportion in Iowa (97.0%) than in NC (91.7%), and most were married or cohabiting (88.5% in Iowa and 83.7% in NC). Significant differences in age, race and ethnicity, marital status, and educational attainment were observed between the two states (p < 0.05).

**Table 1.**
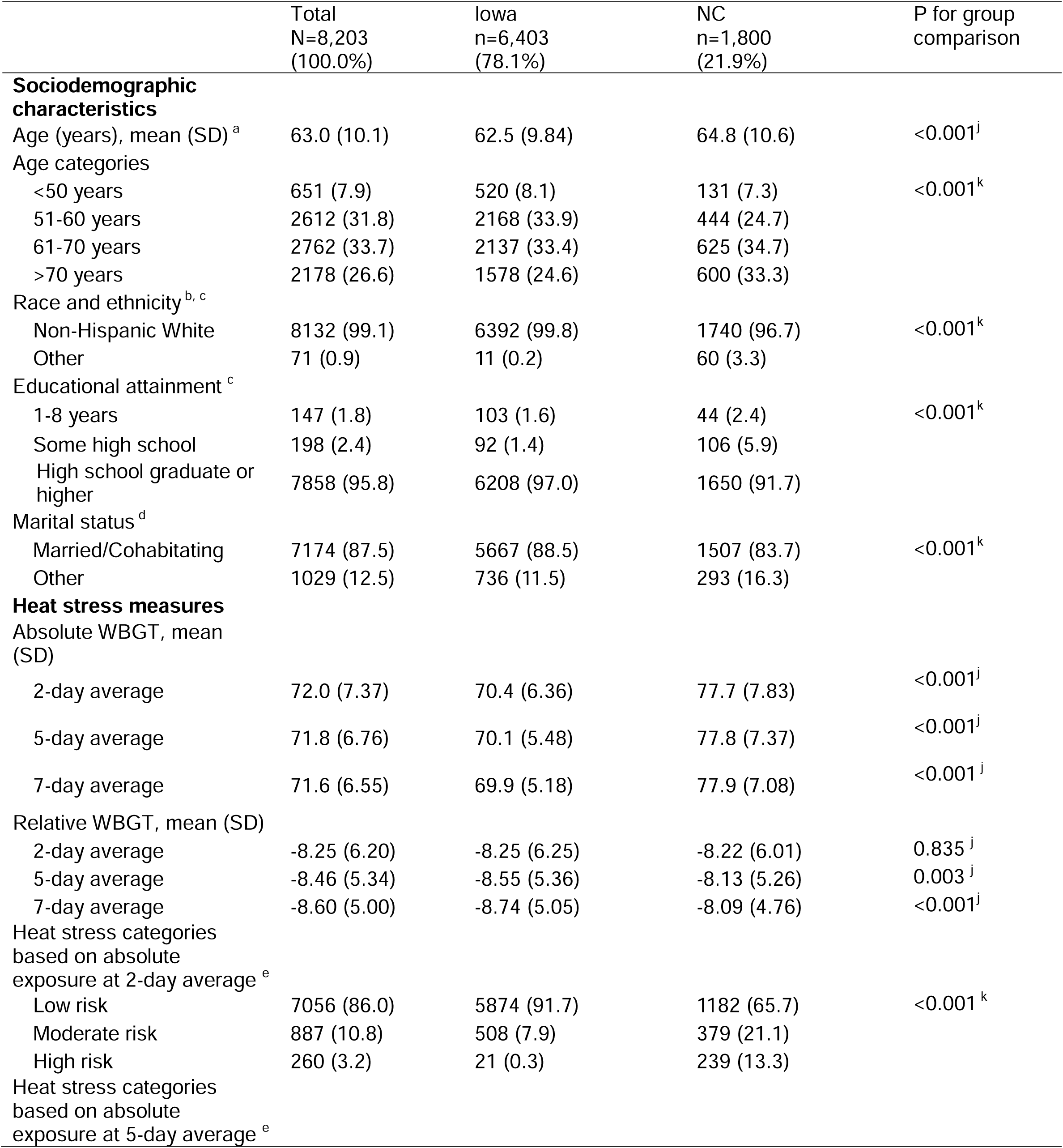
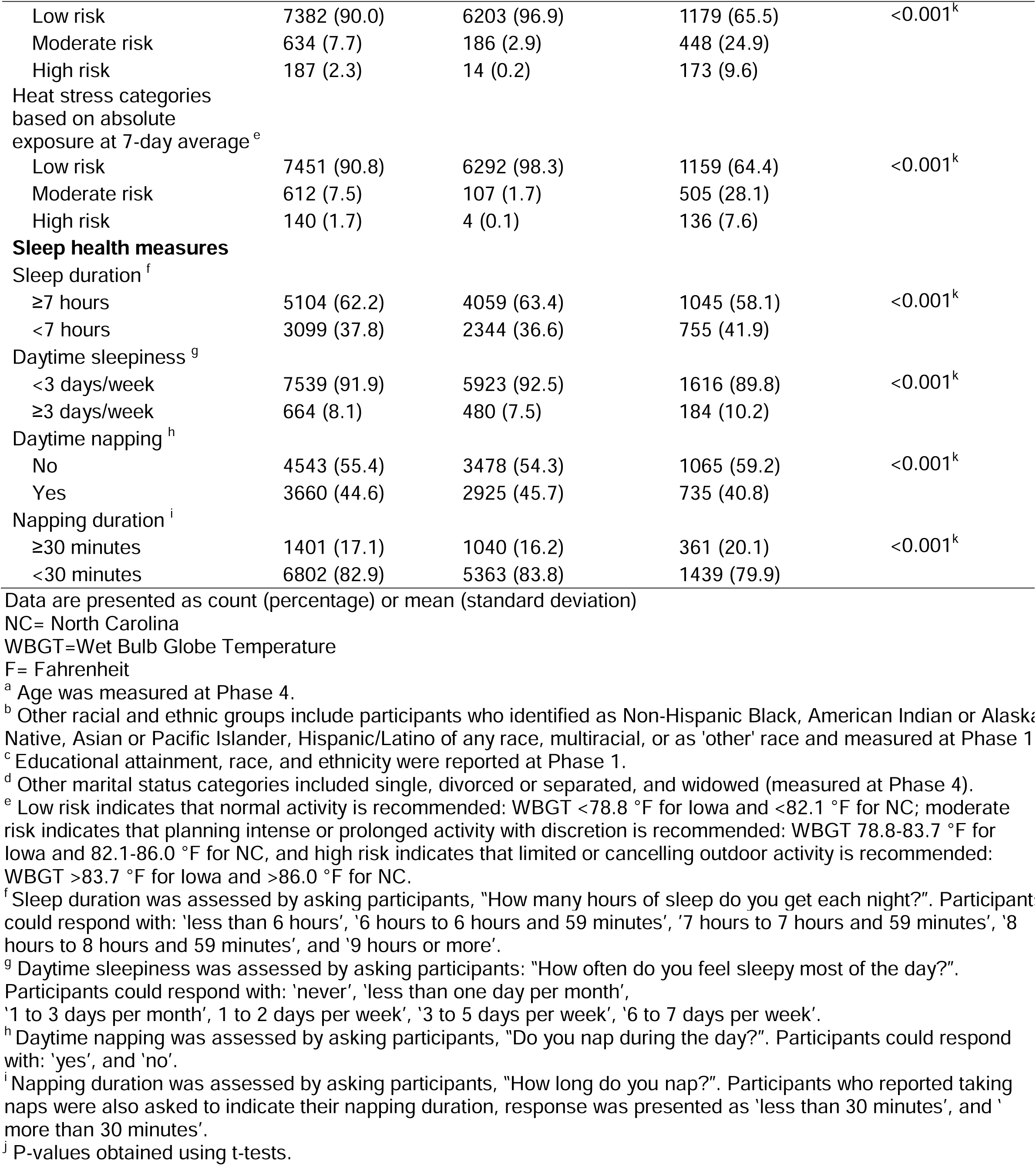
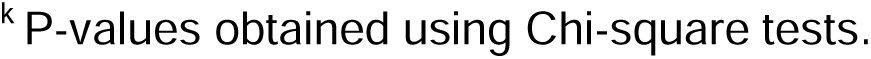
Sociodemographic characteristics, sleep health, and heat stress measures among male farmers, 2013-2015.

|  | Total<br>N=8,203<br>(100.0%) | Iowa<br>n=6,403<br>(78.1%) | NC<br>n=1,800<br>(21.9%) | P for group<br>comparison |
| --- | --- | --- | --- | --- |
| **Sociodemographic characteristics** |  |  |  |  |
| Age (years), mean (SD) <sup>a</sup> | 63.0 (10.1) | 62.5 (9.84) | 64.8 (10.6) | <0.001<sup>j</sup> |
| Age categories |  |  |  |  |
| <50 years | 651 (7.9) | 520 (8.1) | 131 (7.3) | <0.001<sup>k</sup> |
| 51-60 years | 2612 (31.8) | 2168 (33.9) | 444 (24.7) |  |
| 61-70 years | 2762 (33.7) | 2137 (33.4) | 625 (34.7) |  |
| >70 years | 2178 (26.6) | 1578 (24.6) | 600 (33.3) |  |
| Race and ethnicity <sup>b, c</sup> |  |  |  |  |
| Non-Hispanic White | 8132 (99.1) | 6392 (99.8) | 1740 (96.7) | <0.001<sup>k</sup> |
| Other | 71 (0.9) | 11 (0.2) | 60 (3.3) |  |
| Educational attainment <sup>c</sup> |  |  |  |  |
| 1-8 years | 147 (1.8) | 103 (1.6) | 44 (2.4) | <0.001<sup>k</sup> |
| Some high school | 198 (2.4) | 92 (1.4) | 106 (5.9) |  |
| High school graduate or higher | 7858 (95.8) | 6208 (97.0) | 1650 (91.7) |  |
| Marital status <sup>d</sup> |  |  |  |  |
| Married/Cohabiting | 7174 (87.5) | 5667 (88.5) | 1507 (83.7) | <0.001<sup>k</sup> |
| Other | 1029 (12.5) | 736 (11.5) | 293 (16.3) |  |
| **Heat stress measures** |  |  |  |  |
| Absolute WBGT, mean (SD) |  |  |  |  |
| 2-day average | 72.0 (7.37) | 70.4 (6.36) | 77.7 (7.83) | <0.001<sup>j</sup> |
| 5-day average | 71.8 (6.76) | 70.1 (5.48) | 77.8 (7.37) | <0.001<sup>j</sup> |
| 7-day average | 71.6 (6.55) | 69.9 (5.18) | 77.9 (7.08) | <0.001<sup>j</sup> |
| Relative WBGT, mean (SD) |  |  |  |  |
| 2-day average | -8.25 (6.20) | -8.25 (6.25) | -8.22 (6.01) | 0.835<sup>j</sup> |
| 5-day average | -8.46 (5.34) | -8.55 (5.36) | -8.13 (5.26) | 0.003<sup>j</sup> |
| 7-day average | -8.60 (5.00) | -8.74 (5.05) | -8.09 (4.76) | <0.001<sup>j</sup> |
| Heat stress categories based on absolute exposure at 2-day average <sup>e</sup> |  |  |  |  |
| Low risk | 7056 (86.0) | 5874 (91.7) | 1182 (65.7) | <0.001<sup>k</sup> |
| Moderate risk | 887 (10.8) | 508 (7.9) | 379 (21.1) |  |
| High risk | 260 (3.2) | 21 (0.3) | 239 (13.3) |  |
| Heat stress categories based on absolute exposure at 5-day average <sup>e</sup> |  |  |  |  |
| Low risk | 7382 (90.0) | 6203 (96.9) | 1179 (65.5) | <0.001<sup>k</sup> |
| Moderate risk | 634 (7.7) | 186 (2.9) | 448 (24.9) |  |
| High risk | 187 (2.3) | 14 (0.2) | 173 (9.6) |  |
| Heat stress categories based on absolute exposure at 7-day average <sup>e</sup> |  |  |  |  |
| Low risk | 7451 (90.8) | 6292 (98.3) | 1159 (64.4) | <0.001<sup>k</sup> |
| Moderate risk | 612 (7.5) | 107 (1.7) | 505 (28.1) |  |
| High risk | 140 (1.7) | 4 (0.1) | 136 (7.6) |  |
| **Sleep health measures** |  |  |  |  |
| Sleep duration <sup>f</sup> |  |  |  |  |
| ≥7 hours | 5104 (62.2) | 4059 (63.4) | 1045 (58.1) | <0.001<sup>k</sup> |
| <7 hours | 3099 (37.8) | 2344 (36.6) | 755 (41.9) |  |
| Daytime sleepiness <sup>g</sup> |  |  |  |  |
| <3 days/week | 7539 (91.9) | 5923 (92.5) | 1616 (89.8) | <0.001<sup>k</sup> |
| ≥3 days/week | 664 (8.1) | 480 (7.5) | 184 (10.2) |  |
| Daytime napping <sup>h</sup> |  |  |  |  |
| No | 4543 (55.4) | 3478 (54.3) | 1065 (59.2) | <0.001<sup>k</sup> |
| Yes | 3660 (44.6) | 2925 (45.7) | 735 (40.8) |  |
| Napping duration <sup>i</sup> |  |  |  |  |
| ≥30 minutes | 1401 (17.1) | 1040 (16.2) | 361 (20.1) | <0.001<sup>k</sup> |
| <30 minutes | 6802 (82.9) | 5363 (83.8) | 1439 (79.9) |  |

Data are presented as count (percentage) or mean (standard deviation)
NC= North Carolina
WBGT=Wet Bulb Globe Temperature
F= Fahrenheit
<sup>a</sup> Age was measured at Phase 4.
<sup>b</sup> Other racial and ethnic groups include participants who identified as Non-Hispanic Black, American Indian or Alaska Native, Asian or Pacific Islander, Hispanic/Latino of any race, multiracial, or as 'other' race and measured at Phase 1
<sup>c</sup> Educational attainment, race, and ethnicity were reported at Phase 1.
<sup>d</sup> Other marital status categories included single, divorced or separated, and widowed (measured at Phase 4).
<sup>e</sup> Low risk indicates that normal activity is recommended: WBGT <78.8 °F for Iowa and <82.1 °F for NC; moderate risk indicates that planning intense or prolonged activity with discretion is recommended: WBGT 78.8-83.7 °F for Iowa and 82.1-86.0 °F for NC, and high risk indicates that limited or cancelling outdoor activity is recommended: WBGT >83.7 °F for Iowa and >86.0 °F for NC.
<sup>f</sup> Sleep duration was assessed by asking participants, "How many hours of sleep do you get each night?". Participant could respond with: 'less than 6 hours', '6 hours to 6 hours and 59 minutes', '7 hours to 7 hours and 59 minutes', '8 hours to 8 hours and 59 minutes', and '9 hours or more'.
<sup>g</sup> Daytime sleepiness was assessed by asking participants: "How often do you feel sleepy most of the day?".

WBGT indices differed between Iowa and NC. For example, using the 2-day average exposure, the mean absolute WBGTs were 70.4°F (SD=6.36) in Iowa and 77.7°F (SD=7.83) in NC. The relative WBGT were −8.25 °F (SD=6.25) in Iowa and −8.22 °F (SD=6.01) in NC. Iowa experienced more ‘normal activity’ or low risk of heat stress days than in NC (e.g., 91.7% vs. 65.7%). Regarding sleep outcomes, 36.6% in Iowa and 41.9% of participants in NC reported short sleep duration. Daytime sleepiness was reported by 7.5% in Iowa and 10.2% in NC.

Daytime napping was reported by 45.7% in Iowa and 40.8% in NC, while 16.2% in Iowa and 20.1% in NC napped for ≥30 minutes.

### Associations between heat stress and sleep outcomes

Table 2 presents absolute and relative WBGT in relation to sleep duration, daytime sleepiness, napping, and napping duration in both Iowa and NC. Two-day, 5-day, and 7-day average absolute WBGT were not significantly associated with short sleep duration (e.g., PR_Iowa_ = 1.01 [95% CI: 1.00–1.01] and PR_NC_ = 1.00 [95% CI: 0.99–1.02] for 2-day average absolute WBGT) nor daytime sleepiness (e.g., PR_Iowa_ = 1.00 [95% CI: 0.99–1.00] and PR_NC_ = 0.99 [95% CI: 0.98–1.01] for 2-day average absolute WBGT). There were no associations between per 0.1 SD increase in absolute or relative WBGT and daytime napping or longer napping duration in Iowa. However, in NC, absolute WBGT was associated with a higher likelihood of daytime napping for 2-day average exposure (PR_NC_ = 1.02 [95% CI: 1.01–1.04]), for 5-day average exposure (PR_NC_ = 1.02 [95% CI: 1.01–1.04]) and for 7-day average exposure (PR_NC_ = 1.02 [95% CI: 1.00–1.03]). A consistent finding was observed when using relative 2-day WBGT (PR_NC_ = 1.02 [95% CI: 1.00–1.04]) in NC. No associations were observed between absolute or relative WBGT and napping duration in NC.

**Table 2.** The association between each 1 SD increase in WBGT and sleep health among male farmers, PR (95%CI).

|  |  | PR (95%CI) |  |  |  |  |  |
| --- | --- | --- | --- | --- | --- | --- | --- |
|  |  | Iowa (n=6,403) |  |  | NC (n=1,800) |  |  |
|  |  | 2-day WBGT | 5-day WBGT | 7-day WBGT | 2-day WBGT | 5-day WBGT | 7-day WBGT |
| Sleep duration |  |  |  |  |  |  |  |
| Absolute |  | 1.01 (1.00-1.01) | 1.00 (1.00-1.01) | 1.01 (1.00-1.01) | 1.00 (0.99-1.02) | 1.01 (0.99-1.02) | 1.01 (0.99-1.02) |
| Relative |  | 1.01 (1.00-1.01) | 1.01 (1.00-1.01) | 1.01 (1.00-1.02) | 1.01 (0.99-1.02) | 1.01 (0.99-1.02) | 1.01 (0.99-1.02) |
| Daytime sleepiness |  |  |  |  |  |  |  |
| Absolute |  | 1.00 (0.99-1.00) | 1.00 (0.99-1.00) | 1.00 (0.99-1.00) | 0.99 (0.98-1.01) | 1.00 (0.98-1.01) | 1.00 (0.98-1.01) |
| Relative |  | 0.99 (0.99-1.00) | 0.99 (0.99-1.00) | 1.00 (0.99-1.00) | 1.00 (0.98-1.01) | 1.00 (0.99-1.01) | 1.00 (0.98-1.01) |
| Daytime napping |  |  |  |  |  |  |  |
| Absolute |  | 1.01 (1.00-1.01) | 1.01 (1.00-1.02) | 1.01 (1.00-1.01) | **1.02 (1.01-1.04)** | **1.02 (1.01-1.04)** | **1.02 (1.00-1.03)** |
| Relative |  | 1.00 (1.00-1.01) | 1.01 (1.00-1.01) | 1.01 (1.00-1.01) | **1.02 (1.00-1.04)** | 1.01 (1.00-1.03) | 1.01 (1.00-1.03) |
| Napping duration |  |  |  |  |  |  |  |
| Absolute |  | 0.99 (0.99-1.00) | 1.00 (0.99-1.00) | 1.00 (0.99-1.01) | 1.00 (0.99-1.02) | 1.00 (0.99-1.02) | 1.00 (0.99-1.02) |
| Relative |  | 0.99 (0.98-1.00) | 1.00 (0.99-1.00) | 1.00 (0.99-1.00) | 1.00 (0.99-1.02) | 1.00 (0.99-1.01) | 1.00 (0.99-1.01) |

SD: standard deviation; NC: North Carolina; CI: confidence interval; PR: prevalence ratio; WBGT: Wet Bulb Globe Temperature All models were adjusted for age, marital status, educational attainment, and race and ethnicity. Boldface indicates statistical significance at a two-sided p-value of 0.05.

We found associations between WBGT categories and sleep duration, daytime napping, and napping duration (Table 3). First, in Iowa, moderate risk for heat stress, measured by the 2-day WBGT average, was associated with a higher prevalence of short sleep duration (PR= 1.04 [95% CI: 1.00–1.07]). Second, moderate and high risk for heat stress, as assessed by the 2-day average WBGT, was associated with a higher likelihood of daytime napping (e.g., PR_NC_ = 1.06 [95% CI: 1.02–1.11] for high risks) in NC. Moderate risk for heat stress, as assessed by the 7-day average WBGT, was also associated with a higher likelihood of daytime napping (PR_NC_ = 1.04 [95% CI: 1.00–1.07] for moderate risks). Third, in both NC and Iowa, high risk for heat stress across multiple WBGT exposure periods was associated with a lower prevalence of a napping duration of ≥30 minutes (e.g., PR_Iowa, 2-day WBGT_ = 0.86 [95% CI: 0.83–0.89] and PR_NC, 5-day WBGT_ = 0.95 [95% CI: 0.91–1.00]).

**Table 3.** The association between categories of WBGT a and sleep health among male farmers, PR (95%CI).

|  |  | PR (95%CI) |  |  |  |  |  |
| --- | --- | --- | --- | --- | --- | --- | --- |
|  |  | Iowa (n=6,403) |  |  | NC (n=1,800) |  |  |
|  |  | 2-day WBGT | 5-day WBGT | 7-day WBGT | 2-day WBGT | 5-day WBGT | 7-day WBGT |
| Sleep duration |  |  |  |  |  |  |  |
| Moderate risk |  | **1.04 (1.00-1.07)** | 1.03 (0.98-1.08) | 1.01 (0.94-1.08) | 1.01 (0.97-1.05) | 1.00 (0.96-1.04) | 1.02 (0.98-1.06) |
| High risk |  | 1.06 (0.91-1.22) | 1.04 (0.87-1.24) | 0.91 (0.65-1.27) | 1.01 (0.96-1.05) | 1.04 (0.99-1.10) | 1.04 (0.98-1.10) |
| Daytime sleepiness |  |  |  |  |  |  |  |
| Moderate risk |  | 1.01 (0.99-1.03) | 0.99 (0.96-1.02) | 0.98 (0.94-1.02) | 0.99 (0.96-1.02) | 0.99 (0.96-1.02) | 0.99 (0.96-1.02) |
| High risk |  | 0.97 (0.89-1.06) | 1.00 (0.88-1.13) | 1.16 (0.82-1.64) | 0.99 (0.95-1.03) | 1.01 (0.97-1.06) | 0.98 (0.94-1.03) |
| Daytime napping |  |  |  |  |  |  |  |
| Moderate risk |  | 1.01 (0.98-1.04) | 1.02 (0.97-1.07) | 1.00 (0.94-1.07) | **1.04 (1.00-1.08)** | 1.03 (0.99-1.07) | **1.04 (1.00-1.07)** |
| High risk |  | 0.95 (0.83-1.10) | 1.03 (0.86-1.23) | 1.26 (0.95-1.67) | **1.06 (1.02-1.11)** | 1.02 (0.96-1.07) | 1.01 (0.95-1.07) |
| Napping duration |  |  |  |  |  |  |  |
| Moderate risk |  | 1.00 (0.97-1.02) | 1.00 (0.96-1.05) | 1.02 (0.96-1.08) | 1.01 (0.97-1.05) | 1.01 (0.97-1.05) | 1.02 (0.98-1.05) |
| High risk |  | **0.86 (0.83-0.89)** | **0.89 (0.86-0.92)** | **0.89 (0.86-0.93)** | 1.02 (0.98-1.07) | **0.95 (0.91-1.00)** | 0.96 (0.91-1.01) |

SD: standard definition; NC: North Carolina; CI: confidence interval; PR: prevalence ratio; WBGT: Wet Bulb Globe Temperature
All models were adjusted for age, marital status, educational attainment, and race and ethnicity.
Boldface indicates statistical significance at a two-sided p-value of 0.05.
<sup>a</sup> Low risk indicates that normal activity is recommended: WBGT <78.8 °F for Iowa and <82.1 °F for NC; moderate risk indicates that planning intense or prolonged activity with discretion is recommended: WBGT 78.8-83.7 °F for Iowa and 82.1-86.0 °F for NC, and high risk indicates that limited or cancelling outdoor activity is recommended: WBGT >83.7 °F for Iowa and >86.0 °F for NC.

Age did not modify the associations between WBGT indicators and sleep health among farmers in Iowa (Table 4) and NC (Table 5) (P-values of test for interaction between age and WBGT indicators > 0.05).

**Table 4.** The association between each 1 SD increase in WBGT and sleep health across age groups in Iowa, PR (95%CI).

|  |  | PR (95%CI) |  |  |  |  |  |
| --- | --- | --- | --- | --- | --- | --- | --- |
|  |  | Age <60 years (n=2,688) |  |  | Age ≥60 years (n=3,715) |  |  |
|  |  | 2-day WBGT | 5-day WBGT | 7-day WBGT | 2-day WBGT | 5-day WBGT | 7-day WBGT |
| Sleep duration |  |  |  |  |  |  |  |
| Absolute |  | 1.00 (0.99-1.02) | 1.00 (0.99-1.02) | 1.00 (0.99-1.01) | 1.01 (1.00-1.02) | 1.01 (1.00-1.02) | 1.01 (1.00-1.02) |
| Relative |  | 1.00 (0.99-1.02) | 1.00 (0.99-1.01) | 1.00 (0.99-1.01) | 1.01 (1.00-1.02) | 1.01 (1.00-1.02) | **1.01 (1.00-1.02)** |
| Daytime sleepiness |  |  |  |  |  |  |  |
| Absolute |  | 0.99 (0.98-1.00) | 0.99 (0.98-1.00) | 0.99 (0.98-1.00) | 1.00 (0.99-1.01) | 1.00 (0.99-1.01) | 1.00 (0.99-1.01) |
| Relative |  | 0.99 (0.98-1.00) | 0.99 (0.99-1.00) | 1.00 (0.99-1.00) | 1.00 (0.99-1.00) | 0.99 (0.99-1.00) | 1.00 (0.99-1.00) |
| Daytime napping |  |  |  |  |  |  |  |
| Absolute |  | 1.00 (0.99-1.01) | 1.00 (0.99-1.01) | 1.00 (0.99-1.01) | **1.01 (1.00-1.02)** | **1.01 (1.00-1.02)** | **1.01 (1.00-1.02)** |
| Relative |  | 1.00 (0.99-1.01) | 1.00 (0.99-1.01) | 1.00 (0.99-1.01) | 1.01 (1.00-1.02) | 1.01 (1.00-1.02) | 1.01 (1.00-1.02) |
| Napping duration |  |  |  |  |  |  |  |
| Absolute |  | 0.99 (0.98-1.01) | 1.00 (0.99-1.01) | 1.00 (0.99-1.01) | 0.99 (0.98-1.00) | 1.00 (0.98-1.01) | 1.00 (0.99-1.01) |
| Relative |  | 1.00 (0.99-1.01) | 1.00 (0.99-1.01) | 1.00 (0.99-1.01) | 0.99 (0.98-1.00) | 0.99 (0.98-1.00) | 1.00 (0.98-1.01) |

SD: standard definition; NC: North Carolina; CI: confidence interval; PR: prevalence ratio; WBGT: Wet Bulb Globe Temperature
All models were adjusted for age, marital status, educational attainment, and race and ethnicity.
Boldface indicates statistical significance at a two-sided p-value of 0.05.
All P-values of test for interaction were >0.05.

**Table 5.** The association between each 1 SD increase in WBGT and sleep health across age groups in NC, PR (95%CI).

|  |  | PR (95%CI) |  |  |  |  |  |
| --- | --- | --- | --- | --- | --- | --- | --- |
|  |  | Age <60 years (n=575) |  |  | Age ≥60 years (n=1,225) |  |  |
|  |  | 2-day WBGT | 5-day WBGT | 7-day WBGT | 2-day WBGT | 5-day WBGT | 7-day WBGT |
| Sleep duration |  |  |  |  |  |  |  |
|  | Absolute | 1.01 (0.98-1.03) | 1.01 (0.98-1.03) | 1.01 (0.98-1.04) | 1.00 (0.98-1.02) | 1.00 (0.98-1.02) | 1.00 (0.98-1.02) |
|  | Relative | 1.00 (0.98-1.03) | 1.01 (0.98-1.03) | 1.01 (0.98-1.04) | 1.01 (0.99-1.03) | 1.01 (0.99-1.03) | 1.01 (0.99-1.03) |
| Daytime sleepiness |  |  |  |  |  |  |  |
|  | Absolute | 1.00 (0.98-1.02) | 1.00 (0.98-1.02) | 1.00 (0.98-1.02) | 0.99 (0.98-1.01) | 1.00 (0.98-1.01) | 0.99 (0.98-1.01) |
|  | Relative | 1.00 (0.98-1.02) | 1.00 (0.98-1.02) | 1.00 (0.98-1.02) | 0.99 (0.98-1.01) | 1.00 (0.98-1.01) | 1.00 (0.98-1.01) |
| Daytime napping |  |  |  |  |  |  |  |
|  | Absolute | 1.01 (0.99-1.04) | 1.02 (0.99-1.04) | 1.01 (0.98-1.04) | **1.03 (1.01-1.05)** | **1.02 (1.00-1.04)** | **1.02 (1.00-1.04)** |
|  | Relative | 1.01 (0.98-1.04) | 1.01 (0.98-1.04) | 1.01 (0.98-1.03) | **1.03 (1.01-1.04)** | 1.02 (1.00-1.04) | 1.01 (0.99-1.03) |
| Napping duration |  |  |  |  |  |  |  |
|  | Absolute | 1.00 (0.97-1.02) | 1.00 (0.98-1.02) | 1.00 (0.98-1.02) | 1.01 (0.99-1.03) | 1.01 (0.99-1.03) | 1.01 (0.99-1.02) |
|  | Relative | 1.00 (0.98-1.02) | 1.01 (0.99-1.03) | 1.01 (0.99-1.03) | 1.00 (0.98-1.02) | 1.00 (0.98-1.02) | 0.99 (0.98-1.01) |

NC: North Carolina; CI: confidence interval; PR: prevalence ratio; WBGT: Wet Bulb Globe Temperature
All models were adjusted for age, marital status, educational attainment, and race and ethnicity.
Boldface indicates statistical significance at a two-sided p-value of 0.05.
All P-values of test for interaction were >0.05.

### Sensitivity analysis

In the sensitivity analysis, when female farmers were included in the analysis, the main findings—such as the association between 2-day average WBGT and napping—did not change substantially (Tables S2–S3). After changing the cut point for defining short sleep duration (Table S4-S5), the association between moderate risk WBGT exposure, measured by the 2-day average, and sleep duration <8 hours in Iowa was consistent with the main findings (PR = 1.06 [95% CI: 1.01-1.10], see Table S5). Changing the thresholds for relative WBGT also did not affect our findings substantially (Table S6). For example, the significant relationship between 2-day relative WBGT exposure measured by 90^th^ (PR = 1.02 [95% CI: 1.01-1.04]) and 95^th^ percentiles (PR = 1.02 [95% CI: 1.00-1.03]) with daytime napping in NC were consistent with the main results using the initial cut point (i.e., 92.5^th^ percentile). In the models examining the association between heat stress and sleep duration, adjusting for daytime napping did not change our findings. Consistent results were also observed in models assessing the impact of heat stress on daytime napping after adjusting for sleep duration (Table S7).

## DISCUSSION

In our investigation of heat stress in relation to sleep health among farmers in Iowa and North Carolina, heat stress was associated with poor sleep, highlighting its potential impact on nighttime sleep duration, daytime napping, and napping duration. These findings were generally consistent with our hypothesis. In particular, we observed that short-term exposure to heat stress was associated with a higher likelihood of daytime napping in NC. In Iowa, WBGT exceeding the moderate threshold for risk of heat stress was associated with short nightly sleep duration. High risk of heat stress was associated with a lower likelihood of daytime napping 30 minutes or more in both states. We did not observe effect modification by age, which was inconsistent with our hypothesis.

The influence of temperature on sleep health is emerging as a critical public health issue ^11, 13, 60, 61^. Most previous studies enrolled samples of the general population rather than outdoor workers such as farmers. These prior studies have also mainly focused on ambient temperature ^15, 17^, such as the heat index ^28^; however, the heat index does not account for factors such as daytime sun exposure, wind, sun angle, or cloud cover ^23^. Our study extends previous research by focusing on farmers who represent a population more exposed than the general population and by using WBGT as a key indicator to assess heat stress. Our results were also comparable to prior literature. For the Iowa sample, moderate risk of heat stress (i.e., 2-day average exposure) was associated with short sleep duration of <7 hours. Similar to this finding of a positive relationship between higher WBGT levels and shorter sleep duration, evidence among general population from one study conducted in northwestern rural of Burkina Faso also reported a positive relationship between nighttime WBGT ≥25°C and shorter sleep duration among 143 participants.^28^ However, unlike our findings, a recent study including 83 participants conducted in Kenya did not observe a significant correlation between nightly minimum WBGT and sleep duration, ^29^ but these differences might be partially explained by the timing and types of the measurement indicators used (e.g., daytime vs. nighttime; average vs. minimum WBGT). Moreover, we only observed a significant association between heat stress and sleep duration among famers in Iowa rather than NC. This may be due to a higher proportion of participants in Iowa being exposed to lower levels of heat stress and, therefore, may have had lower adaptation to heat upon exposure. As a result, when heat stress increased to moderate levels, their sleep duration was more likely to be reduced.

There are several potential explanations for the heat stress and short sleep duration relationship. Heat exposure contributes to increased wakefulness and shorter sleep duration, and humidity further increases wakefulness ^62^. Heat adaptation also contributes to the alterations in sleep. Prior studies highlighted that heat-related sleep disruptions persist after 5 days of continuous daytime and nocturnal heat exposure ^63^. In fact, in our study, the association between moderate risk of heat stress and sleep duration were stronger in Iowa than in NC, despite NC having higher WBGT than Iowa, indicating the need for more attention in regions with less heat adaption. A final point to note is that, in the sensitivity analysis, where short sleep duration was redefined as <8 hours per night, we still observed the adverse association of heat stress on sleep duration in Iowa, with larger magnitudes of association than observed in the main findings. This suggests that the adverse impact of heat stress on sleep duration likely remains, even after potential misreporting or overestimation of sleep duration.

We observed associations between heat stress and higher prevalence of daytime napping, as shown in NC. Although prior studies have reported the impact of heat on nightly sleep ^18, 19, 39^, few have specifically investigated its influence on daytime napping. Through heat acclimatization, the body adjusts to high temperatures by sweating and increasing blood flow to the skin ^64^, yet this process forces the body to expend extra energy. For example, blood vessels dilate to release heat, resulting in lower blood pressure, which can further contribute to fatigue ^65^. Furthermore, napping can mitigate acute stress and improve emotional well-being, which may serve as a compensatory strategy to recover from fatigue caused by daytime heat ^66, 67^.

Previous research has reported that a 40-minute daytime nap after a late evening simulated soccer match improves short-term sports performance in soccer players and has positive effects on the perception of sleepiness ^68^. Thus, outdoor workers may feel fatigued but use napping as a strategy to avoid the heat of the day, recover energy, and cool down after finishing their work. This may also explain the lack of association between WBGT and daytime sleepiness in our study. Additionally, high risk to heat stress exposure was associated with shorter napping duration, as seen in NC and Iowa. Heat stress exposure may lead farmers to take naps, but napping duration does not necessarily increase. Consistent with our study, results from a study in Hungary showed that higher daily mean temperatures were associated with reduced total sleep time (the sum of night and napping durations) ^15^. Another study suggested that the association between heat and sleep stages are concentrated in the initial segment of the sleep cycle, making it more difficult to fall asleep ^69^. Also, while napping may be a strategy to combat fatigue, increased heat stress can hinder heat dissipation, adversely reducing core body cooling and prolonging wakefulness ^70^. These combined observations suggest that heat stress can affect napping duration, leading to shorter and possibly poorer quality naps. However, our data are limited in determining whether napping is a direct result of insufficient sleep duration, which warrants further investigation. Nonetheless, our findings extend previous research by reporting an association between heat stress and daytime napping, contributing to a deeper understanding of how heat stress may affect daytime sleep patterns among outdoor workers.

We did not find evidence that age modified the relationship between heat stress and sleep in either NC or Iowa. One possible explanation is that some older participants—such as those over age 70—did not meet the analysis inclusion and exclusion criteria, as presented in Table S1. The remaining participants may have had a greater ability to adapt to heat stress compared to those more vulnerable to its effects. Additionally, the sample sizes for each age group in NC were relatively small, which may have limited the ability to detect age-related differences. More research is warranted to explore the potential modifying effect of age.

There are several limitations to consider. First, the cross-sectional design of our study does not allow for causal inferences. Second, residential addresses at enrollment were used for linkages to WBGT measurements; misclassification is possible if participants moved. However, farmers are generally more residentially stable, as they tend to live on or near their farms, which could reduce the likelihood of significant exposure misclassification. Third, recall of self-reported sleep was not in reference to a specific time frame (e.g., past 24 hours, past week). Therefore, there may be a disconnect between the timing of the heat exposure metrics and the reference period of the sleep questions, potentially obscuring associations. However, results were consistent across the 3 assessed exposure periods. Fourth, measurement error of sleep is possible ^57^. Subjective measures of sleep were collected via self-administered and computer-assisted telephone interviews, which may introduce potential recall bias. Also, due to limited data, we used daytime sleepiness as a proxy for sleep quality, as poor sleep is known to predict daytime sleepiness. However, this measure does not fully capture the nuances of sleep quality, as daytime sleepiness can be influenced by both sleep quantity and quality ^71^. Fifth, since sleep can be influenced by both daytime and nighttime temperatures, future studies should incorporate measurements of both daytime and nighttime temperatures/heat stress. Sixth, our sample primarily consisted of farmers who were men, underscoring the need for future studies to investigate the impact of heat on outdoor workers who are women. Additionally, there is a need for more research on other racial and ethnic groups, as this study predominantly involved non-Hispanic White farmers. The sample was older on average (like the general farming population), so future studies should ensure the inclusion of younger populations as well, including laborers who may have greater vulnerabilities and fewer opportunities for daytime napping. Our analyses assumed outdoor work, though some farmers may have mitigated their heat exposure by behaviors such as drinking ice water, using air conditioner, tractors with airconditioned cabs, accessing shade, and performing office work; however, these data were unavailable. Measurement and investigation of these factors and their potential impacts on associations are warranted in future studies. Vulnerability to heat stress may be increased with greater physical exertion and obesity ^72^, which we did not assess. Further research is needed with sufficient sample sizes and varying sociodemographic characteristics to improve representation and generalizability. Since the participants were predominantly from two states in the US, the study findings cannot be extrapolated to other regions, which suggests the need for nationally representative samples as well as studies that include other regions. Lastly, some relevant behavioral data were not available such as using air conditioners and drinking ice water which could confound the associations.

Despite the limitations, our study has several strengths. First, we investigated heat stress in relation to multiple sleep outcomes (including daytime and nighttime sleep) among farmers, representing an understudied topic and population in sleep research. Notably, the use of WBGT which accounts for the thermal conditions affecting personal comfort, extends previous literature that primarily focused on assessing ambient temperature in relation to health outcomes including sleep ^15, 17, 18^. Moreover, our sample includes both relative and absolute heat measures with different exposure window periods (2-, 5-, and 7-day); we accounted for both acute exposure and more long-term climate adaptation. Furthermore, the farmers in our study were confirmed to be personally involved in farming activities and residing on farms, though they may not be representative of other outdoor workers. Samples from NC and Iowa, representing two different geographical regions, provide insights into two typical geographical conditions in the US with differing average exposures to heat stress. These strengths provide valuable evidence that improves understanding the potential impact of exposure to heat on sleep health.

## CONCLUSION

Short-term heat stress exposure was associated with a higher likelihood of shorter sleep duration among farmers in Iowa and daytime napping among farmers in NC. Additionally, high risk of heat stress was linked to shorter napping duration among farmers in both states. Future studies should use longitudinal data and cohorts of farmers across the US as well as objective sleep indicators to confirm the heat stress and sleep relationship we observed among farmers. After replication in more generalizable samples of farmers, findings can inform the potential impact of working in extreme heat conditions and guide interventions (e.g., heat stroke or injury prevention/mitigation) related to, for instance, tailoring working hours, shift breaks, and outdoor workload to promote recommended sleep duration and quality sleep among farmers.

## Data Availability Statement

Data are available through approved requests based on Agricultural Health Study policy.

## Code availability

The datasets and code used and/or analyzed during the current study are available from the corresponding author on reasonable request.

## Ethical Approval

The National Institute of Environmental Health Sciences Institutional Review Board approved the Agricultural Health Study protocol. Approval for use of de-identified, secondary data in this analysis is exempt from the United States federal regulations for the protection of human research participants. All participants provided implied consent at enrollment and ongoing active consent during follow-up surveys following protocols approved by the relevant Institutional Review Boards. All methods were performed in accordance with the relevant guidelines and regulations.

## Data Availability

All data produced in the present study are available upon reasonable request to the authors.

## Data Availability

All data produced in the present study are available upon reasonable request to the authors.

## Acknowledgements

The authors wish to thank the AHS participants as well as Aubrey Hubbard, PhD, MPH (at DLH, LLC) for the quality control review of the data analysis.

## Author Contributions Statement

***Authors:*** *WS. Zhou, SA. Gaston, BT. Ogbenna, C. Parks, DP. Sandler, CL. Jackson*.

*Study concept: CL. Jackson*.

*Study design: WS. Zhou, SA. Gaston, CL. Jackson. Acquisition of data: CP. Parks, D. Sandler*.

*Statistical Analysis: WS. Zhou*.

*Interpretation of data: WS. Zhou, SA. Gaston, BT. Ogbenna, CP. Parks, D. Sandler, CL. Jackson*.

*Drafting of the manuscript: WS. Zhou*.

*Critical revision of the manuscript for important intellectual content: WS. Zhou, SA. Gaston, BT. Ogbenna, C. Parks, DP. Sandler, CL. Jackson*.

*Administrative, technical, and material support: SA. Gaston, C. Parks, CL. Jackson. Obtaining funding: CL. Jackson, DP. Sandler*.

*Study supervision: CL. Jackson*.

*Final Approval: WS. Zhou, SA. Gaston, BT. Ogbenna, C. Parks, DP. Sandler, CL. Jackson*

## Supplemental materials

**Figure S1.**
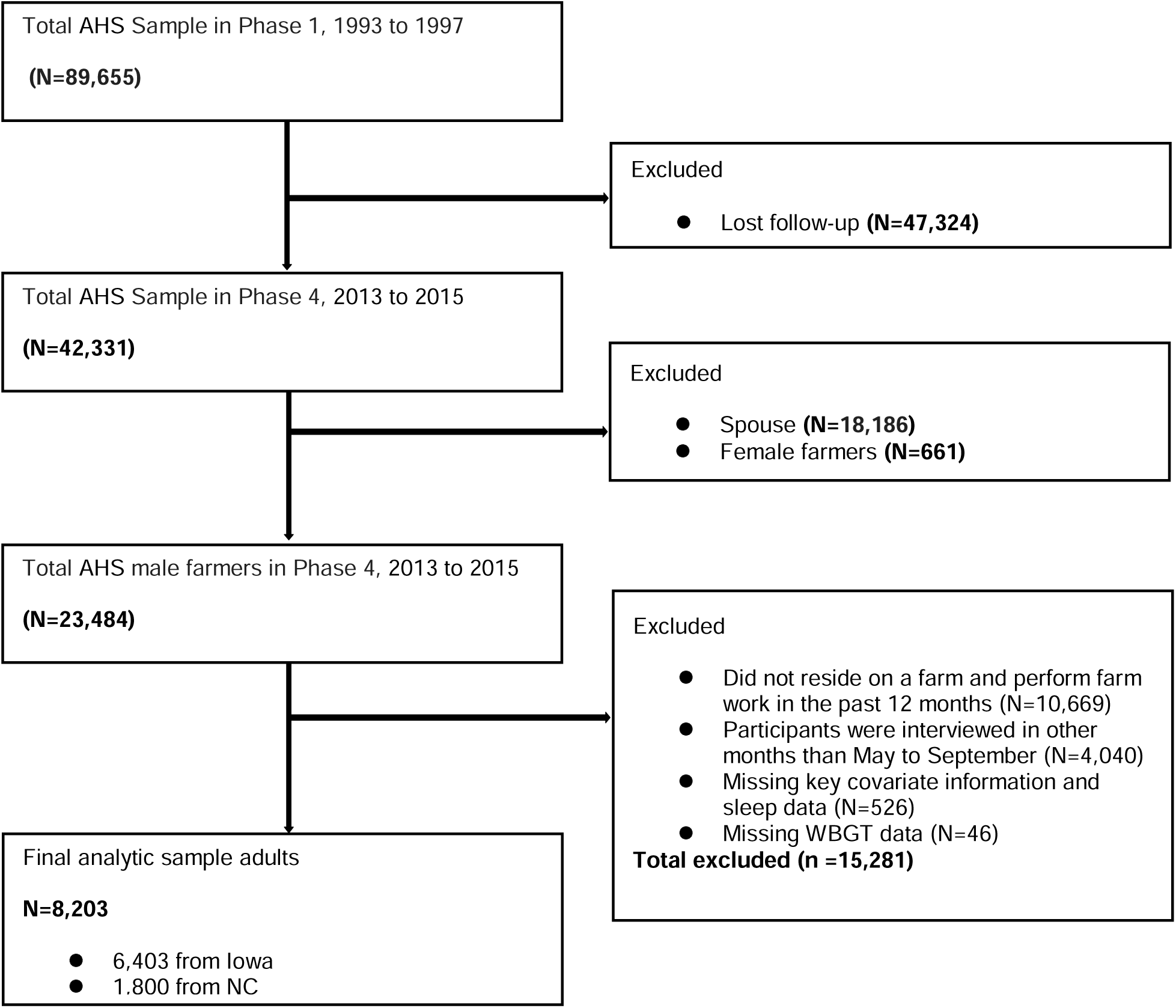
The flow chart of sample selection

**Figure S2.**
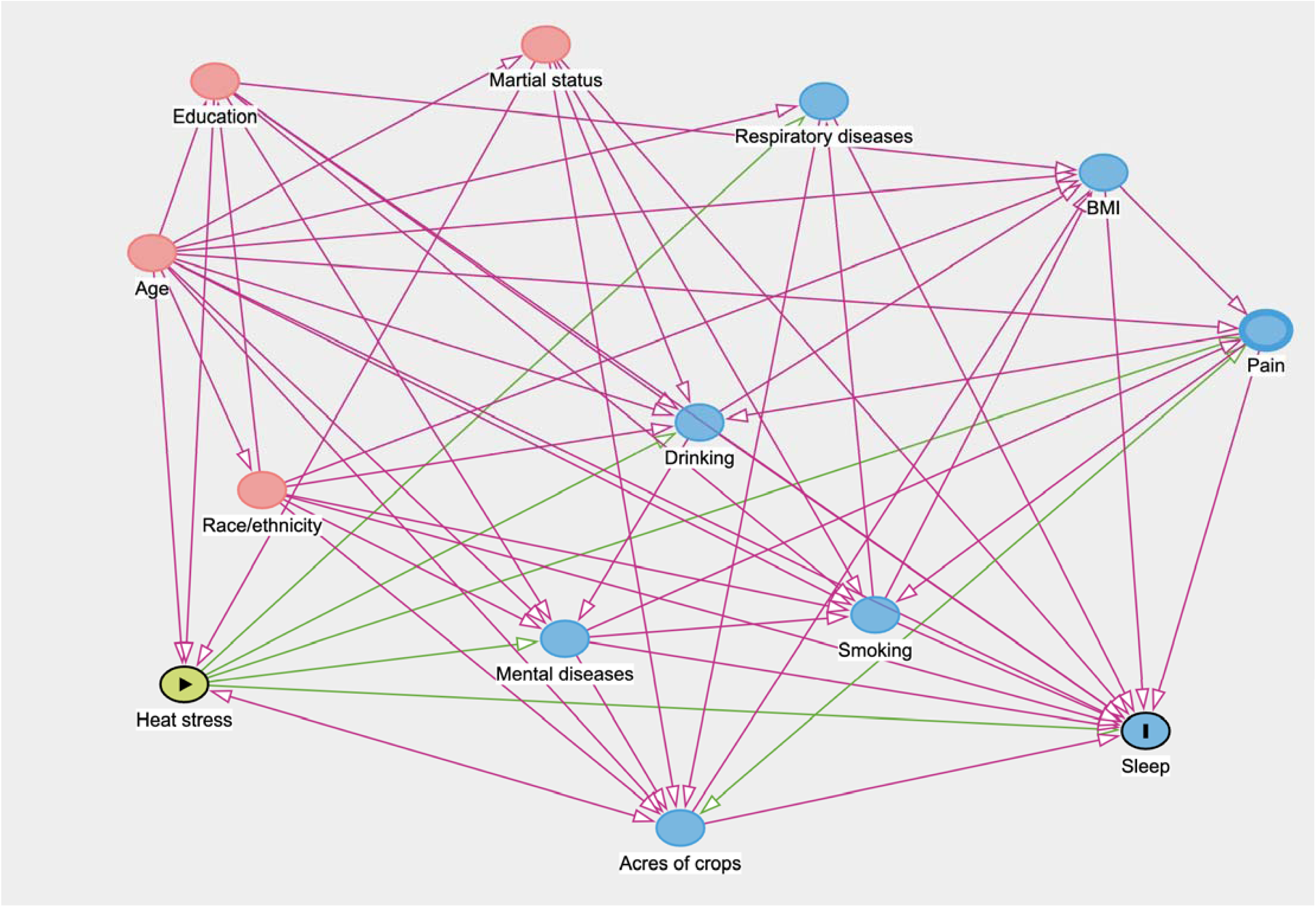
Directed Acyclic Graph (DAG) for the association between heat stress and sleep

**Table S1.** Basic characteristics among excluded and included participants.

|  | Included:<br>n=8,203 | Excluded:<br>n=15,281 |
| --- | --- | --- |
| **Sociodemographic characteristics** |  |  |
| Age, mean (SD) <sup>a</sup> | 63.0 (10.1) | 68.3 (12.1) |
| Missing | 0 | 0 |
| Age categories |  |  |
| <50 | 651 (7.9) | 911 (6.0) |
| 51-60 | 2612 (31.8) | 3019 (19.8) |
| 61-70 | 2762 (33.7) | 4119 (27.0) |
| >70 | 2178 (26.6) | 7232 (47.3) |
| Missing | 0 | 0 |
| Race and ethnicity <sup>b, c</sup> |  |  |
| Non-Hispanic White | 8132 (99.1) | 14636 (95.8) |
| Other | 71 (0.9) | 355 (2.3) |
| Missing | 0 | 290 (1.9) |
| Educational attainment <sup>c</sup> |  |  |
| 1-8 years | 147 (1.8) | 643 (4.2) |
| Some high school | 198 (2.4) | 761 (5.0) |
| High school graduate or higher | 7858 (95.8) | 13059 (85.5) |
| Missing | 0 | 818 (5.4) |
| Marital status <sup>d</sup> |  |  |
| Married/Cohabiting | 7174 (87.5) | 10803 (70.7) |
| Other | 1029 (12.5) | 2007 (13.1) |
| Missing | 0 | 2471 (16.2) |
| Geographical region |  |  |
| Iowa | 6403 (78.1) | 9461 (61.9) |
| NC | 1800 (21.9) | 5668 (37.1) |
| Missing | 0 | 152 (1.0) |
| **Sleep outcomes** |  |  |
| Sleep duration <sup>e</sup> |  |  |
| ≥7 hours | 5104 (62.2) | 7906 (51.7) |
| <7 hours | 3099 (37.8) | 4794 (31.4) |
| Missing | 0 | 2581 (16.9) |
| Daytime sleepiness <sup>f</sup> |  |  |
| <3 days/week | 7539 (91.9) | 11191 (73.2) |
| ≥3 days/week | 664 (8.1) | 1399 (9.2) |
| Missing | 0 | 2691 (17.6) |
| Daytime napping <sup>g</sup> |  |  |
| No | 4543 (55.4) | 6572 (43.0) |
| Yes | 3660 (44.6) | 6130 (40.1) |
| Missing | 0 | 2579 (16.9) |
| Napping duration <sup>h</sup> |  |  |
| ≥30 minutes | 1401 (17.1) | 3145 (20.6) |
| <30 minutes | 6802 (82.9) | 9399 (61.5) |
| Missing | 0 | 2737 (17.9) |

Data are presented as count (percentage) or mean (standard deviation)
NC= North Carolina
<sup>a</sup> Age was measured at Phase 4.
<sup>b</sup> Other racial and ethnic groups include participants who identified as Non-Hispanic Black, American Indian or Alaska Native, Asian or Pacific Islander, Hispanic/Latino of any race, multiracial, or as 'other' race and measured at Phase 1.
<sup>c</sup> Educational attainment, race, and ethnicity were reported at Phase 1.
<sup>d</sup> Other marital status categories included single, divorced or separated, and widowed (measured at Phase 4).
<sup>e</sup> Sleep duration was assessed by asking participants, "How many hours of sleep do you get each night?". Participants could respond with: 'less than 6 hours', '6 hours to 6 hours and 59 minutes', '7 hours to 7 hours and 59 minutes', '8 hours to 8 hours and 59 minutes', and '9 hours or more'.
<sup>f</sup> Daytime sleepiness was assessed by asking participants: "How often do you feel sleepy most of the day?". Participants could respond with: 'never', 'less than one day per month', '1 to 3 days per month', '1 to 2 days per week', '3 to 5 days per week', '6 to 7 days per week'.
<sup>g</sup> Daytime napping was assessed by asking participants, "Do you nap during the day?". Participants could respond with: 'yes', and 'no'.
<sup>h</sup> Napping duration was assessed by asking participants, "How long do you nap?". Participants who reported taking naps were also asked to indicate their napping duration, response was presented as 'less than 30 minutes', and 'more than 30 minutes'.

**Table S2.** The association between each 1 SD increase in WBGT and sleep health among male and female farmers, PR (95%CI).

|  |  | PR (95%CI) |  |  |  |  |  |
| --- | --- | --- | --- | --- | --- | --- | --- |
|  |  | Iowa (n=6,499) |  |  | NC (n=1,854) |  |  |
|  |  | 2-day WBGT | 5-day WBGT | 7-day WBGT | 2-day WBGT | 5-day WBGT | 7-day WBGT |
| Sleep duration |  |  |  |  |  |  |  |
| Absolute |  | 1.00 (1.00-1.01) | 1.00 (0.99-1.01) | 1.01 (1.00-1.01) | 1.00 (0.99-1.02) | 1.00 (0.99-1.02) | 1.00 (0.99-1.02) |
| Relative |  | 1.00 (1.00-1.01) | 1.00 (1.00-1.01) | 1.01 (1.00-1.01) | 1.00 (0.99-1.02) | 1.01 (0.99-1.02) | 1.01 (0.99-1.02) |
| Daytime sleepiness |  |  |  |  |  |  |  |
| Absolute |  | 1.00 (0.99-1.00) | 1.00 (0.99-1.00) | 1.00 (0.99-1.00) | 1.00 (0.98-1.01) | 1.00 (0.99-1.01) | 1.00 (0.98-1.01) |
| Relative |  | 0.99 (0.99-1.00) | 0.99 (0.99-1.00) | 1.00 (0.99-1.00) | 1.00 (0.98-1.01) | 1.00 (0.99-1.01) | 1.00 (0.99-1.01) |
| Daytime napping |  |  |  |  |  |  |  |
| Absolute |  | 1.00 (1.00-1.01) | 1.01 (1.00-1.02) | 1.01 (1.00-1.01) | **1.02 (1.01-1.04)** | **1.02 (1.01-1.04)** | **1.02 (1.00-1.03)** |
| Relative |  | 1.00 (1.00-1.01) | 1.01 (1.00-1.01) | 1.00 (1.00-1.01) | **1.02 (1.00-1.03)** | 1.02 (1.00-1.03) | 1.01 (1.00-1.03) |
| Napping duration |  |  |  |  |  |  |  |
| Absolute |  | 0.99 (0.99-1.00) | 1.00 (0.99-1.00) | 1.00 (0.99-1.01) | 1.00 (0.99-1.02) | 1.00 (0.99-1.02) | 1.00 (0.99-1.02) |
| Relative |  | 0.99 (0.99-1.00) | 1.00 (0.99-1.00) | 1.00 (0.99-1.01) | 1.00 (0.99-1.02) | 1.00 (0.99-1.01) | 1.00 (0.99-1.01) |

SD: standard deviation; NC: North Carolina; CI: confidence interval; PR: prevalence ratio; WBGT: Wet Bulb Globe Temperature All models were adjusted for age, gender, marital status, educational attainment, and race and ethnicity. Boldface indicates statistical significance at a two-sided p-value of 0.05.

**Table S3.** The association between categories of WBGT ^a^ and sleep health among male and female farmers, PR (95%CI).

|  |  | PR (95%CI) |  |  |  |  |  |
| --- | --- | --- | --- | --- | --- | --- | --- |
|  |  | Iowa (n=6,499) |  |  | NC (n=1,854) |  |  |
|  |  | 2-day WBGT | 5-day WBGT | 7-day WBGT | 2-day WBGT | 5-day WBGT | 7-day WBGT |
| Sleep duration |  |  |  |  |  |  |  |
| Moderate risk |  | **1.03 (1.00-1.07)** | 1.03 (0.98-1.08) | 1.01 (0.94-1.08) | 1.00 (0.96-1.04) | 1.00 (0.96-1.03) | 1.02 (0.98-1.05) |
| High risk |  | 1.05 (0.91-1.22) | 1.04 (0.87-1.24) | 0.91 (0.65-1.27) | 1.00 (0.95-1.05) | 1.04 (0.99-1.10) | 1.03 (0.97-1.10) |
| Daytime sleepiness |  |  |  |  |  |  |  |
| Moderate risk |  | 1.01 (0.98-1.03) | 0.99 (0.96-1.02) | 0.98 (0.94-1.02) | 0.99 (0.96-1.02) | 0.99 (0.97-1.02) | 0.99 (0.96-1.02) |
| High risk |  | 0.97 (0.89-1.06) | 1.00 (0.88-1.13) | 1.16 (0.82-1.64) | 0.99 (0.96-1.03) | 1.02 (0.97-1.06) | 0.99 (0.94-1.03) |
| Daytime napping |  |  |  |  |  |  |  |
| Moderate risk |  | 1.01 (0.98-1.04) | 1.02 (0.97-1.07) | 1.00 (0.94-1.07) | **1.04 (1.00-1.08)** | 1.03 (0.99-1.07) | **1.04 (1.00-1.07)** |
| High risk |  | 0.95 (0.83-1.10) | 1.03 (0.86-1.23) | 1.26 (0.95-1.66) | **1.07 (1.02-1.11)** | 1.02 (0.97-1.07) | 1.02 (0.96-1.08) |
| Napping duration |  |  |  |  |  |  |  |
| Moderate risk |  | 1.00 (0.97-1.03) | 1.00 (0.96-1.05) | 1.02 (0.96-1.08) | 1.01 (0.97-1.05) | 1.01 (0.98-1.05) | 1.02 (0.98-1.05) |
| High risk |  | **0.86 (0.83-0.89)** | **0.89 (0.85-0.92)** | **0.89 (0.86-0.92)** | 1.02 (0.98-1.07) | **0.95 (0.91-1.00)** | 0.96 (0.91-1.01) |

NC: North Carolina; CI: confidence interval; PR: prevalence ratio; WBGT: Wet Bulb Globe Temperature All models were adjusted for age, gender, marital status, educational attainment, and race and ethnicity. Boldface indicates statistical significance at a two-sided p-value of 0.05.
<sup>a</sup> Low risk indicates that normal activity is recommended: WBGT <78.8 °F for Iowa and <82.1 °F for NC; moderate risk indicates that planning intense or prolonged activity with discretion is recommended: WBGT 78.8-83.7 °F for Iowa and 82.1-86.0 °F for NC, and high risk indicates that limited or cancelling outdoor activity is recommended: WBGT >83.7 °F for Iowa and >86.0 °F for NC.

**Table S4.**
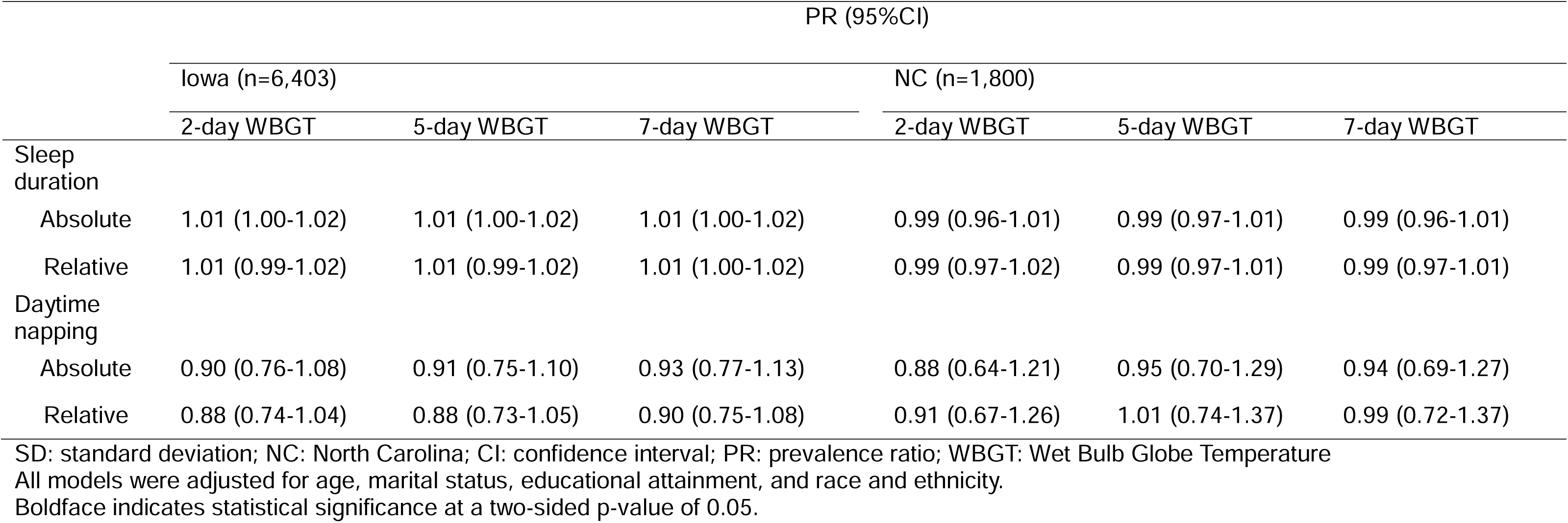
The association between each 1 SD increase in WBGT and sleep health among farmers after changing the cut points for sleep duration (<8 hours vs. **≥** 8 hours) and daytime napping (**≥**1 hour vs. <1 hour), PR (95%CI).

**Table S5.** The association between categories of WBGT ^a^ and sleep health among farmers after changing the cut points for sleep duration (<8 hours vs. **≥** 8 hours) and daytime napping (**≥**1 hour vs. <1 hour), PR (95%CI).

|  |  | PR (95%CI) |  |  |  |  |  |
| --- | --- | --- | --- | --- | --- | --- | --- |
|  |  | Iowa (n=6,403) |  |  | NC (n=1,800) |  |  |
|  |  | 2-day WBGT | 5-day WBGT | 7-day WBGT | 2-day WBGT | 5-day WBGT | 7-day WBGT |
| Sleep duration |  |  |  |  |  |  |  |
| Moderate risk |  | **1.06 (1.01-1.10)** | 1.04 (0.98-1.11) | 1.04 (0.96-1.13) | 0.99 (0.93-1.05) | 0.98 (0.93-1.04) | 1.00 (0.95-1.06) |
| High risk |  | 0.90 (0.70-1.18) | 0.88 (0.64-1.21) | 0.92 (0.53-1.59) | 0.97 (0.91-1.05) | 1.00 (0.93-1.08) | 0.98 (0.89-1.07) |
| Daytime napping |  |  |  |  |  |  |  |
| Moderate risk |  | 1.01 (0.51-2.01) | 0.97 (0.31-3.04) | 1.15 (0.29-4.62) | 0.72 (0.31-1.69) | 1.07 (0.52-2.20) | 1.06 (0.53-2.12) |
| High risk |  | —* | —* | —* | 0.90 (0.34-2.38) | 0.51 (0.12-2.15) | 0.67 (0.16-2.80) |

SD: standard deviation; NC: North Carolina; CI: confidence interval; PR: prevalence ratio; WBGT: Wet Bulb Globe Temperature
<sup>a</sup> Low risk indicates that normal activity is recommended: WBGT <78.8 °F for Iowa and <82.1 °F for NC; moderate risk indicates that planning intense or prolonged activity with discretion is recommended: WBGT 78.8-83.7 °F for Iowa and 82.1-86.0 °F for NC, and high risk indicates that limited or cancelling outdoor activity is recommended: WBGT >83.7 °F for Iowa and >86.0 °F for NC.
All models were adjusted for age, marital status, educational attainment, and race and ethnicity.
Boldface indicates statistical significance at a two-sided p-value of 0.05.
\* Not estimable due to small sample size.

**Table S6.**
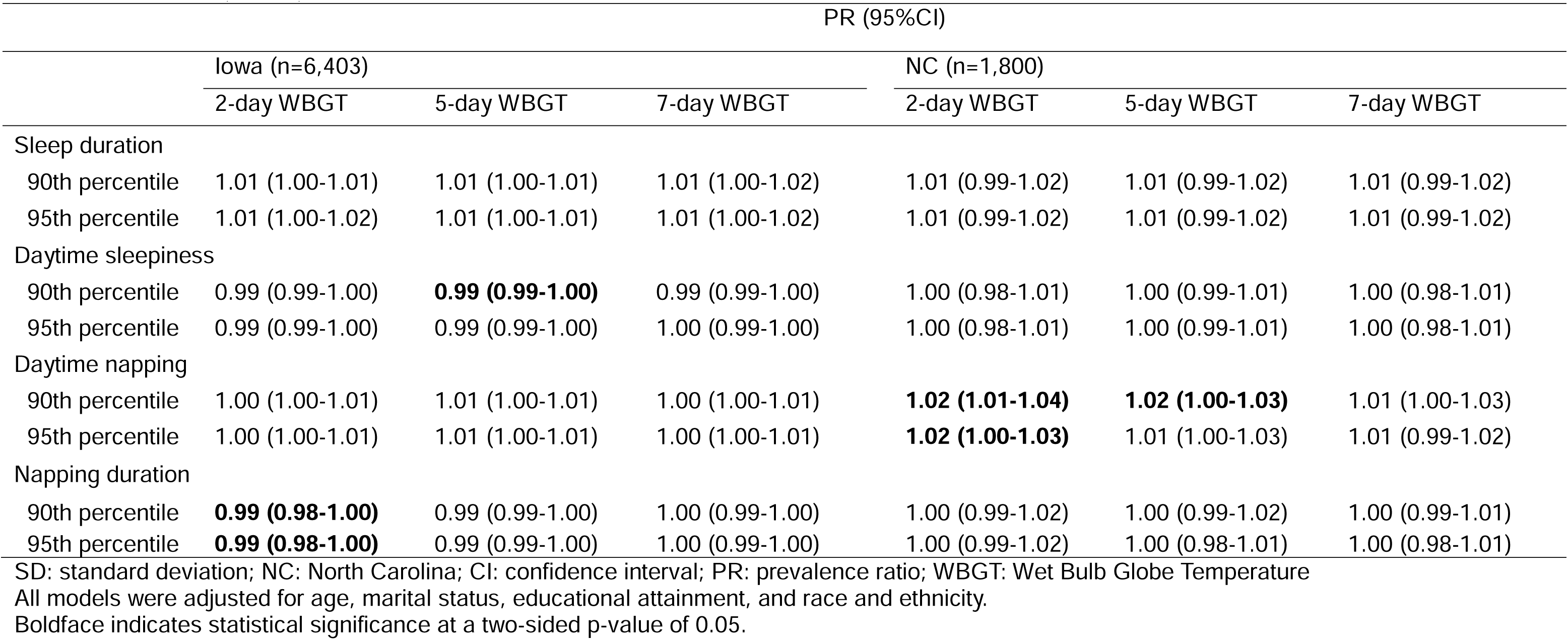
The association between each 1 SD increase in relative heat stress and sleep health among farmers after changing the thresholds for relative WBGT, PR (95%CI).

**Table S7.** The association between each 1 SD increase in WBGT and sleep health among male farmers after adjusting for sleep duration or daytime napping, PR (95%CI).

|  |  | PR (95%CI) |  |  |  |  |  |
| --- | --- | --- | --- | --- | --- | --- | --- |
|  |  | Iowa (n=6,403) |  |  | NC (n=1,800) |  |  |
|  |  | 2-day WBGT | 5-day WBGT | 7-day WBGT | 2-day WBGT | 5-day WBGT | 7-day WBGT |
| Sleep duration <sup>a</sup> |  |  |  |  |  |  |  |
| Absolute |  | 1.00 (1.00-1.01) | 1.00 (1.00-1.01) | 1.01 (1.00-1.02) | 1.00 (0.99-1.02) | 1.01 (0.99-1.02) | 1.01 (0.99-1.02) |
| Relative |  | 1.01 (1.00-1.01) | 1.01 (1.00-1.01) | 1.01 (1.00-1.02) | 1.00 (0.99-1.02) | 1.01 (0.99-1.02) | 1.01 (0.99-1.02) |
| Daytime napping <sup>b</sup> |  |  |  |  |  |  |  |
| Absolute |  | 1.00 (1.00-1.01) | 1.01 (1.00-1.02) | 1.01 (1.00-1.01) | **1.02 (1.01-1.04)** | **1.02 (1.01-1.04)** | **1.02 (1.00-1.03)** |
| Relative |  | 1.00 (1.00-1.01) | 1.01 (1.00-1.01) | 1.00 (1.00-1.01) | **1.02 (1.00-1.04)** | 1.01 (1.00-1.03) | 1.01 (1.00-1.03) |

SD: standard deviation; NC: North Carolina; CI: confidence interval; PR: prevalence ratio; WBGT: Wet Bulb Globe Temperature
<sup>a</sup> Models were adjusted for age, gender, marital status, educational attainment, race and ethnicity, and daytime napping.
<sup>b</sup> Models were adjusted for age, gender, marital status, educational attainment, race and ethnicity, and sleep duration.
Boldface indicates statistical significance at a two-sided p-value of 0.05.

